# Effectiveness and use of evidence-based cardiovascular preventive therapies in type 2 diabetes patients with established or high risk of atherosclerotic cardiovascular disease

**DOI:** 10.1101/2025.02.10.25321971

**Authors:** Jian-Qing Tian, Yu-Hao Lin, Zhi-Jun Zhang, Yi-Ting Peng, Jia-Wen Ye, Zhi-Yi Wang

## Abstract

**Aim:** To explore the association of evidence-based cardiovascular preventive therapies with cardiovascular and renal outcomes in type 2 diabetes (T2DM) patients with established or high risk of atherosclerotic cardiovascular disease (ASCVD).

**Methods:** In this cohort study, we identified T2DM patients with established or high risk of ASCVD using diagnostic codes from the institutional data of Xiamen Humanity Hospital between 2018 and 2023. Cohort 1 includes participants who were visited between 2018 and 2020, with follow-up until occurrence of an endpoint or December 31, 2020. Participants who were visited between 2018 and 2023 were included in cohort 2. A total of 5,335 patients were included in cohort 1, and 17,320 patients were included in cohort 2. Primary outcomes were hazard ratios (HRs) for the composite of 3-point major adverse cardiovascular event (3-P MACE), hospitalization for heart failure (HHF), and end-stage kidney disease or doubling of serum creatinine level.

**Results:** Relative to patients’ non-use of evidence-based cardiovascular preventive therapies, the use of at least one evidence-based cardiovascular preventive therapy was associated with a lower risk of the 3-P MACE (HR, 0.82; 95% confidence interval [CI], 0.67 to 0.98), HHF (HR, 0.66; 95% CI, 0.47 to 0.92) and end-stage kidney disease or doubling of the serum creatinine level (HR, 0.73; 95% CI, 0.60 to 0.89) after adjustment for potential confounders. From 2018 to 2023, the use of glucagon-like peptide 1 receptor agonists increased from 2.7% to 13.7%; sodium–glucose cotransporter 2 inhibitors increased from 3.9% to 16.5%; angiotensin-converting enzyme inhibitors/angiotensin-II receptor blockers increased from 28.1% to 43.0%; moderate-intensity statins increased from 61.6% to 70.5%; and aspirin increased from 23.7% to 32.9%.

**Conclusions:** This study demonstrated that T2DM patients with established or high risk of ASCVD might benefit from the use of evidence-based cardiovascular preventive medications with respect to the risk of 3-P MACE, HHF, and end-stage kidney disease or doubling of the serum creatinine level. Despite a modest annual increase in the use of evidence-based cardiovascular preventive medications in T2DM individuals with established or high risk of ASCVD, multiple strategies are needed to overcome barriers to the implementation of evidence-based therapies.

## 1 Introduction

Type 2 diabetes (T2DM) is a major risk factor for the development of atherosclerotic cardiovascular disease (ASCVD), which is the leading cause of death and long-term complications in T2DM [1]. A systematic review demonstrated that the prevalence of ASCVD in patients with T2DM was nearly 30%, and up to two-thirds of T2DM patients will develop ASCVD during their lifetimes [2,3]. Therefore, there is an urgent need to initiate and intensify secondary preventive to reduce the risk of recurrent cardiovascular events and cardiovascular mortality in patients with T2DM and ASCVD.

Although, the US Food and Drug Administration (FDA) conducted randomized cardiovascular outcome trials (CVOTs) for all glucose–lowering medications in 2008, the cardiovascular benefit of these medications was controversial until 2015. The glucagon-like peptide 1 receptor agonists (GLP-1 RA) and sodium–glucose cotransporter 2 inhibitors (SGLT2i) have shown benefit toward for secondary prevention of ASCVD [4]. Most importantly, large CVOTs of GLP-1 RA and SGLT2i have shown that these drug classes substantially decrease the risk of cardiovascular and renal outcomes [5–11]. What’s more, numerous studies have proved the beneficial effects of angiotensin-converting enzyme inhibitors (ACEIs) or angiotensin-II receptor blockers (ARBs), statins, and aspirin therapy on cardiovascular and renal outcomes [12–14]. Given these accumulating evidences, the American Diabetes Association (ADA) and the Chinese Diabetes Society recommends the cardio-protective glucose-lowering drug classes (GLP-1 RA and SGLT2i), ACEI/ARB, statins, and aspirin for comprehensive cardiovascular risk reduction in adults with T2DM and established or high risk of ASCVD [15,16].

Currently, limited evidence is available for the direct comparison of those using versus not using evidence-based cardiovascular preventive therapies (including: GLP-1 RA, SGLT2i, ACEI/ARB, statins, and aspirin) in terms of cardiovascular, hospitalization for heart failure and renal outcomes. Despite these guidelines recommend the GLP-1 RA, SGLT2i, ACEI/ARB, statins, and aspirin for their cardioprotective effects, a degree of therapeutic inertia in clinical practice may be observed for a variety of reasons.

We therefore compared the cardiovascular and renal benefits of use to non-use evidence-based cardiovascular preventive therapies in adults with T2DM and established or high risk of ASCVD. Accurately determining the gaps in evidence-based therapy is essential to disseminate and promote clinical application of guidelines. Thus, we also describe the use of either GLP-1 RA and/or SGLT2i, ACEI/ARB, moderate-intensity statins, and aspirin in T2DM individuals with established or high risk of ASCVD during the period 2018 to 2023.

## 2 Materials and Methods

### 2.1 Study design and data source

This study was a population-based cohort study. The source population consisted of all outpatients who visited in the Xiamen Humanity Hospital, Fujian Medical University, between January 1, 2018, and December 31, 2023. Informed consent was waived because the study used anonymized patient data. The study protocol was approved by the Xiamen Humanity Hospital Ethics Committee (no. HAXM-MEC-20240 820-048-01). The study was conducted according to the ethical principles of the Declaration of Helsinki. The trial is registered with Chinese Clinical Trial Registry (ChiCTR) (registration no. ChiCTR2500101328).

### 2.2 Study population

The study population included adults (age ≥ 18 years) between January 1, 2018, and December 31, 2023. Data analysis was performed from January 1, 2024, to December 31, 2024. Disease information was identified using the International Statistical Classification of Diseases, Tenth Revision (ICD-10) (eTable 1). ICD-10 diagnosis codes were utilized to identify T2DM patients at high cardiovascular risk, which include patients with concomitant ASCVD or high risk of ASCVD. Briefly, ASCVD was defined as having a history of coronary artery disease (previous prior myocardial infarction, unstable angina), peripheral arterial disease (amputation due to poor circulation), cerebrovascular disease (stroke, or transient ischemic attack), or any revascularization intervention. Patients at high risk of ASCVD were defined as those aged ≥ 55 years with two or more risk factors. Risk factors included obesity (body mass index [BMI] ≥ 28 kg/m2), hypertension, smoking, dyslipidemia, and albuminuria [17]. In addition, age, height, weight, and history of smoking were obtained from an electronic medical record. History of hypertension and dyslipidemia were identified by the presence of ICD-10 diagnosis codes. Albuminuria was defined as urine albumin-to-creatinine ratio ≥ 30 mg/g. Patients younger than 30 at the time of initial insulin prescription, with end-stage kidney disease, or limited life expectancy were excluded.

This study population was divided into two cohorts. Participants who were visited between January 1, 2018, and December 31, 2020, were included in cohort 1 (Figure 1). To reduce the effects of prior treatment, we excluded those who had received medication adjustments in the three months preceding enrollment. Participants were followed from 1 day after cohort entry until occurrence of an endpoint or December 31, 2020. Participants were excluded if they discontinued the index drug, died of other causes, or were lost to follow-up. Cohort 1 was divided into group A (non-use of evidence-based cardiovascular preventive therapies) and group B (use at least one evidence-based cardiovascular preventive therapy). Participants who were visited between January 1, 2018, and December 31, 2023, were included in cohort 2 (Figure 2). Patients with longitudinal follow-up may be included in multiple years but only once per year. Within a given year, only patients with at least 1 glucose-lowering medication record were included. If a patient had multiple visits within a specific year, only the last visit was included in the analysis.

**Figure 1.**
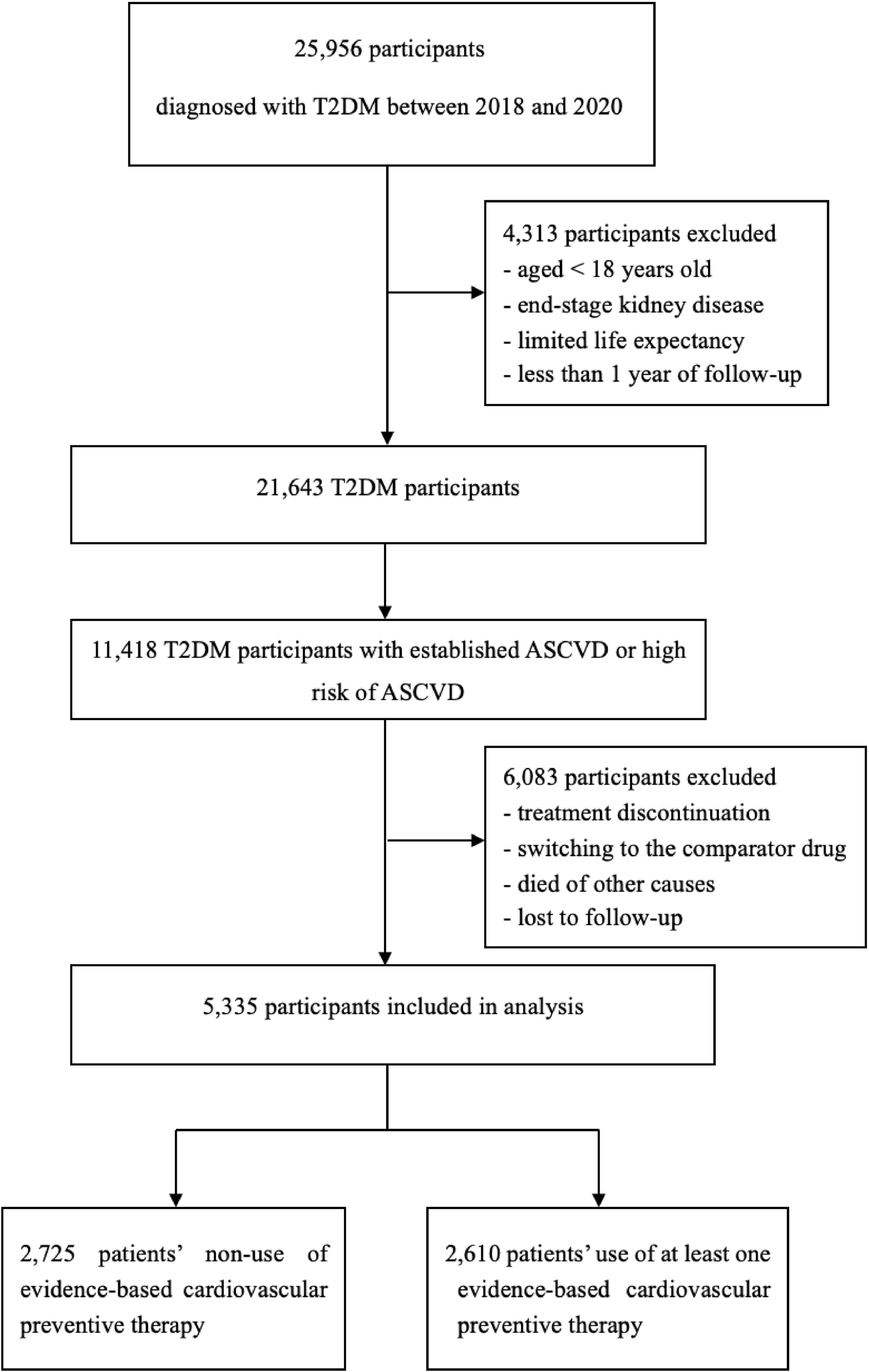
Flow Diagram for Cohort 1. Notes: T2DM, type 2 diabetes mellitus; ASCVD, atherosclerotic cardiovascular disease.

**Figure 2.**
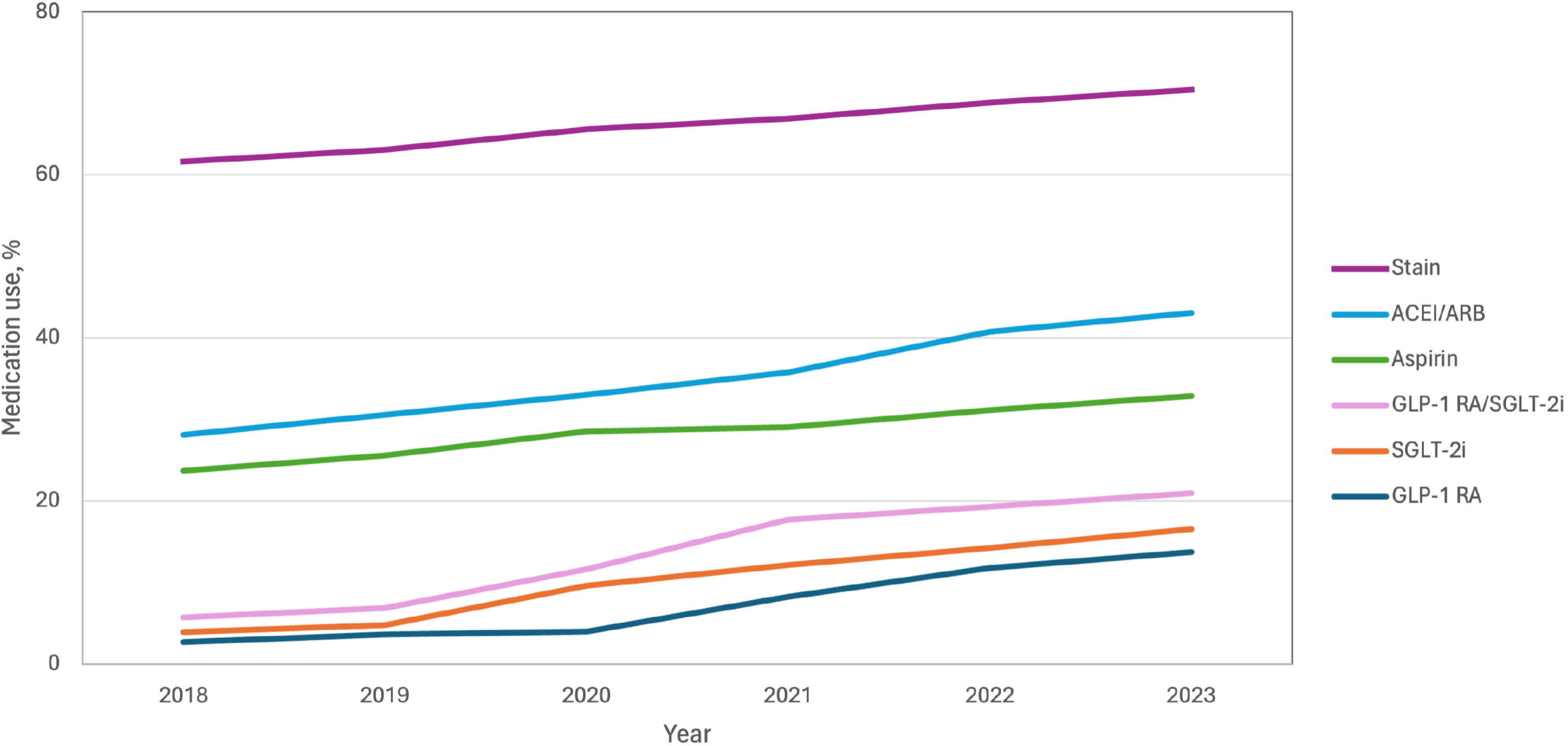
Flow Diagram for Cohort 2. Notes: T2DM, type 2 diabetes mellitus; ASCVD, atherosclerotic cardiovascular disease.

### 2.3 Study Outcomes

We ascertained that the primary outcome was a composite of 3-point major adverse cardiovascular event (cardiovascular death, nonfatal myocardial infarction, or nonfatal stroke), hospitalization for heart failure and end-stage kidney disease or doubling of serum creatinine level in cohort 1. Secondary outcomes that were planned in cohort 1 and cohort 2 were specified in the following order: the difference in clinical characteristics between the use and non-use of evidence-based cardiovascular preventive therapies; the use of evidence-based cardiovascular preventive therapies in T2DM patients with established or high risk of ASCVD. The use of these medications was assessed through the day of the last dispensed prescription within a given year. The prescribed cardioprotective glucose-lowering pharmacotherapy included GLP-1 RA (e.g., semaglutide, dulaglutide, liraglutide, and lixisenatide) and SGLT2i (e.g., empagliflozin, dapagliflozin, and canagliflozin). Prescribed blood pressure-lowering pharmacotherapy included ACEI (e.g., captopril, benazepril, and so on), ARB (e.g., valsartan, irbesartan, and so on), and statins (atorvastatin, rosuvastatin, pitavastatin, simvastatin, and pravastatin).

### 2.4 Covariates

Demographic characteristics, such as age and sex were described. Clinical characteristics included glycosylated hemoglobin (HbA1c), low-density lipoprotein cholesterol (LDL-C), and BMI. For patients with multiple measurements within a given year, only the last one was used. The co-morbidities (heart failure, atrial fibrillation, hypertension, dyslipidemia, and nonalcoholic fatty liver disease) and diabetes chronic complications (retinopathy, neuropathy, and nephropathy) were identified using the ICD-10 code. Concomitant use of oral anti-diabetic, antihypertensive, and lipid-lowering medications was assessed.

### 2.5 Composite Score for Evidence-based Therapy

Evidence-based therapy was defined as the use of either a GLP-1 RA and/or SGLT2i, either an ACEI or ARB, a moderate-intensity statins, and an aspirin. Although high-dose statins are important to prevent cardiovascular events, it is intolerable for most of Chinese populations. Therefore, moderate-intensity statins are recommended as the first choice lipid-lowering drugs by the Endocrinology and Metabolism Physician Branch of the Chinese Medical Doctor Association [18]. Patients in this cohort were deemed to meet the criteria for all four components and were assigned a composite score ranging from 0 to 4, indicating the number of evidence-based therapies prescribed.

### 2.6 Statistical Analysis

Demographic and clinical characteristics, co-morbidities, and concomitant medications were presented as with means ± standard deviations (SDs) for continuous variables and frequency/percentages for categorical variables. The number and proportion of patients with T2DM and established or high risk of ASCVD who received GLP-1 RA and/or SGLT2i, ACEI/ARB, moderate-intensity statins, and aspirin were evaluated annually. Treatment effect was estimated using hazard ratios (HRs) from a Cox regression model. Subgroup analyses were performed by sex, the presence of ASCVD or high risk of ASCVD, the presence of heart failure, and the presence of chronic kidney disease. All data where necessary were conducted to calculate 95% confidence interval (CI). Statistical comparisons baseline characteristics between the different groups were using the independent-sample t test for continuous variables and χ2 test or Fisher’s exact test for categorical variables. Two-sided P values <□0.05 were considered statistically significant. All analyses were performed using R version 4.4.2 (R Foundation).

## 3. Results

### 3.1 Study Participants

We identified 5,335 patients in cohort 1 (2,793 patients with a dual diagnosis of T2DM and ASCVD and 2,524 patients with T2DM and high risk of ASCVD) and 17,320 patients in cohort 2 (8,418 patients with a dual diagnosis of T2DM and ASCVD and 8,902 patients with T2DM and high risk of ASCVD).

Among 5,335 patients in cohort 1, a total of 2,275 patients were assigned to the group A and 2,610 were assigned to the group B. Compared with the group A, patients in the group B were younger (mean [SD]: 65.4 [10.1] years vs 66.8 [10.1] years), more likely to be male (57.5% vs 55.2%), with a lower proportion of second-generation sulfonylurea use (22.4% vs 30.3%) and more common with Urban employee medical insurance (25.8% vs 24.7%). There were differences between group A and group B in clinical characteristics (e.g., 2,275 patients in group A vs 2,610 patients in group B: initiation of SGLT-2i or GLP-1 RAs, 1.6% vs 3.1%; coronary artery disease, 71.2% vs 74.6%; cerebrovascular disease, 20.1% vs 23.3%; heart failure, 30.6% vs 35.8%; CKD stage 3, 18.7% vs 21.5%; mean [SD] Charlson Comorbidity Index score, 4.0 [2.7] vs 4.4 [2.9]) (Table 1).

**Table 1.**
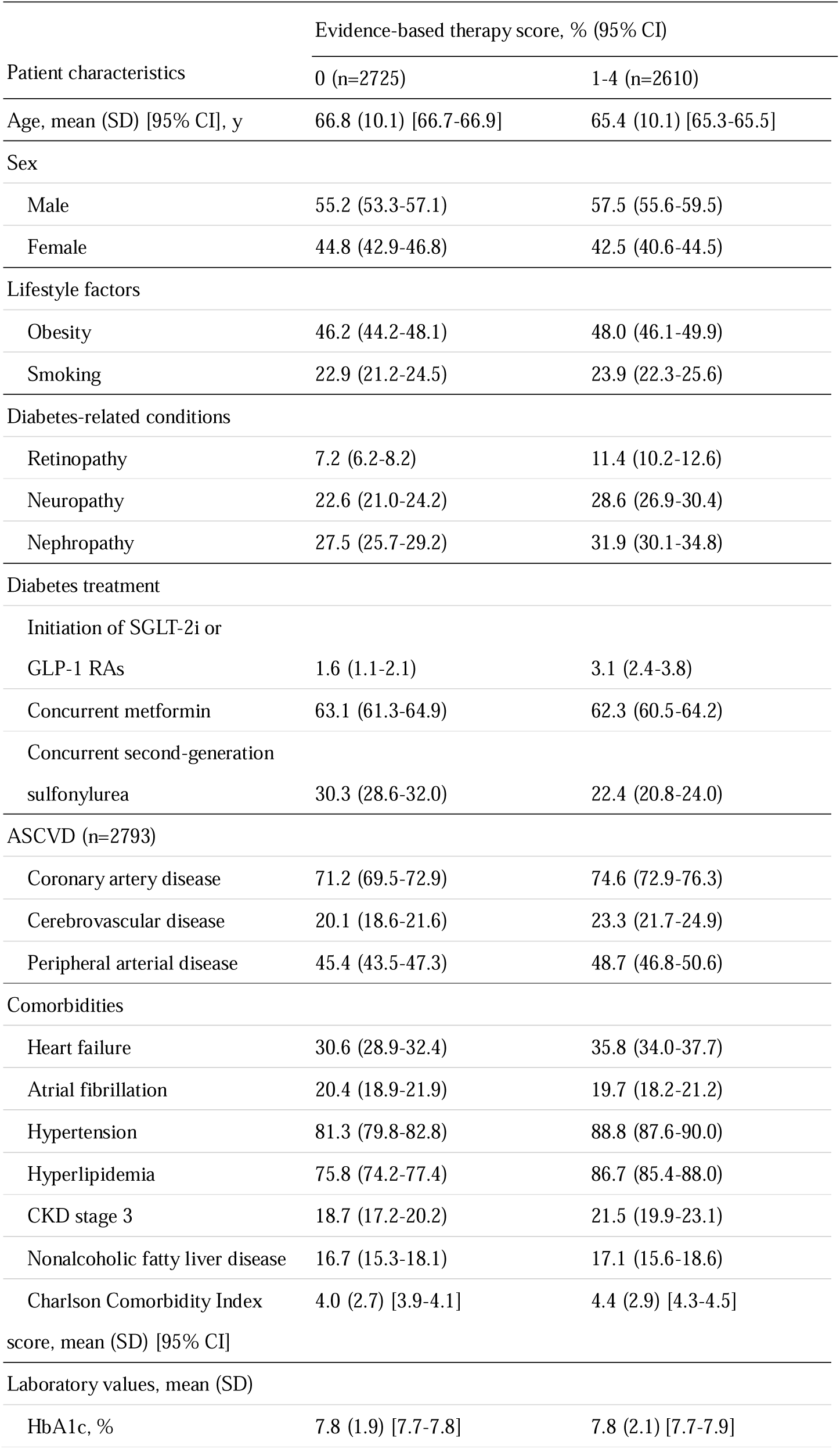
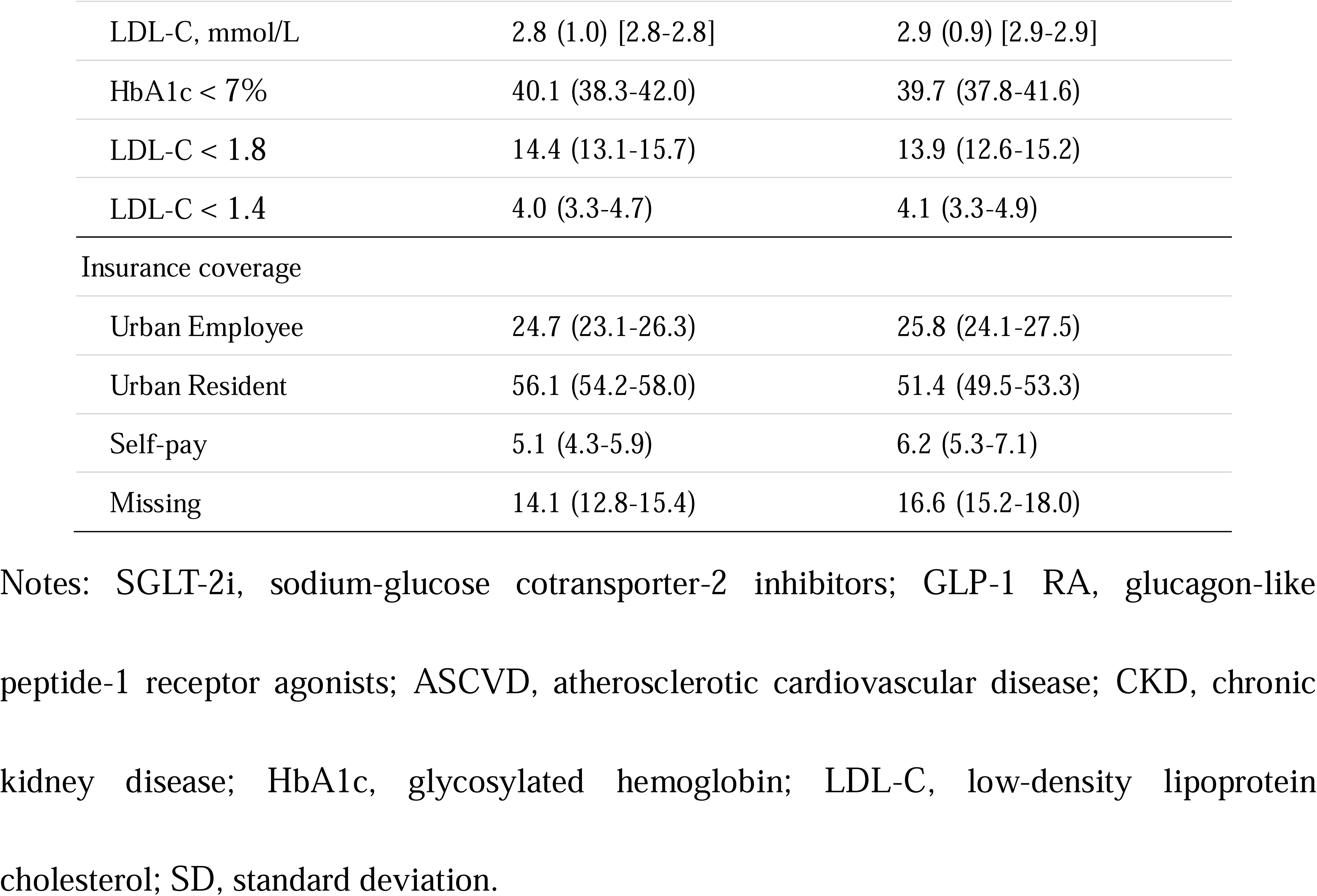
Patient characteristics by overall evidence-based composite score (Cohort 1).

### 3.2 Primary Outcomes

A 3-point major adverse cardiovascular event occurred in 228 of 2,610 patients (8.7%) in the group B (use of at least one evidence-based cardiovascular preventive therapy) and in 237 of 2725 patients (8.7%) in the group A (non-use of evidence-based cardiovascular preventive therapies) (hazard ratio, 0.96; 95% confidence interval [CI], 0.80 to 1.16; P=0.69). Hospitalization for heart failure occurred in 79 of 2,610 patients (3.0%) in the group B and in 103 of 2,725 patients (3.8%) in the group A (hazard ratio, 0.84; 95% CI, 0.62 to 1.12; P=0.23). With respect to the end-stage kidney disease or doubling of the serum creatinine level, the hazard ratio (group B vs group A) was 0.90 (95% CI, 0.75 to 1.09) (Table 2). After adjustment for potential confounders, patients in the group B showed a significant decreased risk of 3-P MACE (hazard ratio, 0.82; 95% CI, 0.67 to 0.98; P=0.036), HHF (hazard ratio, 0.66; 95% CI, 0.47 to 0.92; P<0.01), and end-stage kidney disease or doubling of the serum creatinine level (hazard ratio, 0.73; 95% CI, 0.60 to 0.89; P<0.01) compared with the group A (Table 2). Subgroup analyses did not show a difference in the risk of 3-point MACE, HHF and end-stage kidney disease or doubling of serum creatinine level between the two groups (eTable 2).

**Table 2.** Treatment effect estimates for evidence-based therapies.

| Outcome | Total No. of events (%) |  | Unadjusted |  | Model 1 |  |
| --- | --- | --- | --- | --- | --- | --- |
|  |  |  | Group B vs Group A |  |  |  |
|  | Group B<br>(n=2610) | Group A<br>(n=2725) | HR (95% CI) | *p* | HR (95% CI) | *p* |
| 3-P MACE | 228 (8.7) | 237 (8.7) | 0.96 (0.80-1.16) | 0.69 | 0.82 (0.67-0.98) | 0.036 |
| HHF | 79 (3.0) | 103 (3.8) | 0.84 (0.62-1.12) | 0.23 | 0.66 (0.47-0.92) | <0.01 |
| End-stage kidney disease,<br>doubling of serum creatinine level | 198 (7.6) | 226 (8.3) | 0.90 (0.75-1.09) | 0.29 | 0.73 (0.60-0.89) | <0.01 |

Notes: 3-P MACE, 3-point major adverse cardiovascular event; HHF, hospitalization for heart failure; HR, hazard ratio.
Model 1: Adjusted for sex, age, smoke, BMI, HbA1c, LDL-C, history of diseases including diabetic retinopathy, diabetic nephropathy, diabetic neuropathy, hypertension, and hyperlipidemia.

### 3.3 Use Ratios

Overall, the utilization of glucose-lowering drugs with documented cardiovascular benefits was low; however, the rate increased annually from 2018 to 2023 (GLP-1 RA: from 2.7% [73 of 2,720] to 13.7% [439 of 3,200]; SGLT2i: from 3.9% [107 of 2720] to 16.5% [529 of 3200]). Combined, the percentage of patients with T2DM and established or high risk of ASCVD taking either agent increased from 5.7% (156 of 2,720) in 2018 to 21.0% (671 of 3200) in 2023 (Figure 3). Metformin use was essentially unchanged over this time (61.3% [1667 of 2,720] in 2018 vs 60.8% [1,945 of 3,200] in 2023). Use of noncardiovascular glucose-lowering agents exhibited a variable decline, including alpha-glucosidase inhibitors (30.1% [818 of 2,720] to 22.2% [709 of 3,200]) and dipeptidyl peptidase-4 inhibitors (DPP-4i) (25.5% [694 of 2,720] to 20.8% [665 of 3,200]), although these medications remained more commonly used than SGLT2i or GLP-1 RAs through 2023 (eFigure 1). Of the antihypertensive medications, the use of ACEI/ARB was increased from 28.1% [764 of 2,720] to 43.0% [1,377 of 3,200], while the use of calcium channel blockers was decreased (44.2% [1,202 of 2,720] to 41.8% [1,336 of 3,200]) from 2018 to 2023 (eFigure 2). The use of moderate-intensity statins (61.6% [1,676 of 2,720] to 70.5% [2,255 of 3,200]) and aspirin (23.7% [645 of 2,720] to 32.9% [1,053 of 3,200]) pharmacotherapy variably increased (eFigure 3 and Fig 3). In patients with high risk of ASCVD, GLP-1 RA, SGLT2i, ACEI/ARB, moderate-intensity statins, and aspirin showed lower usage rates than patients with ASCVD (eFigure 4).

**Figure 3.**
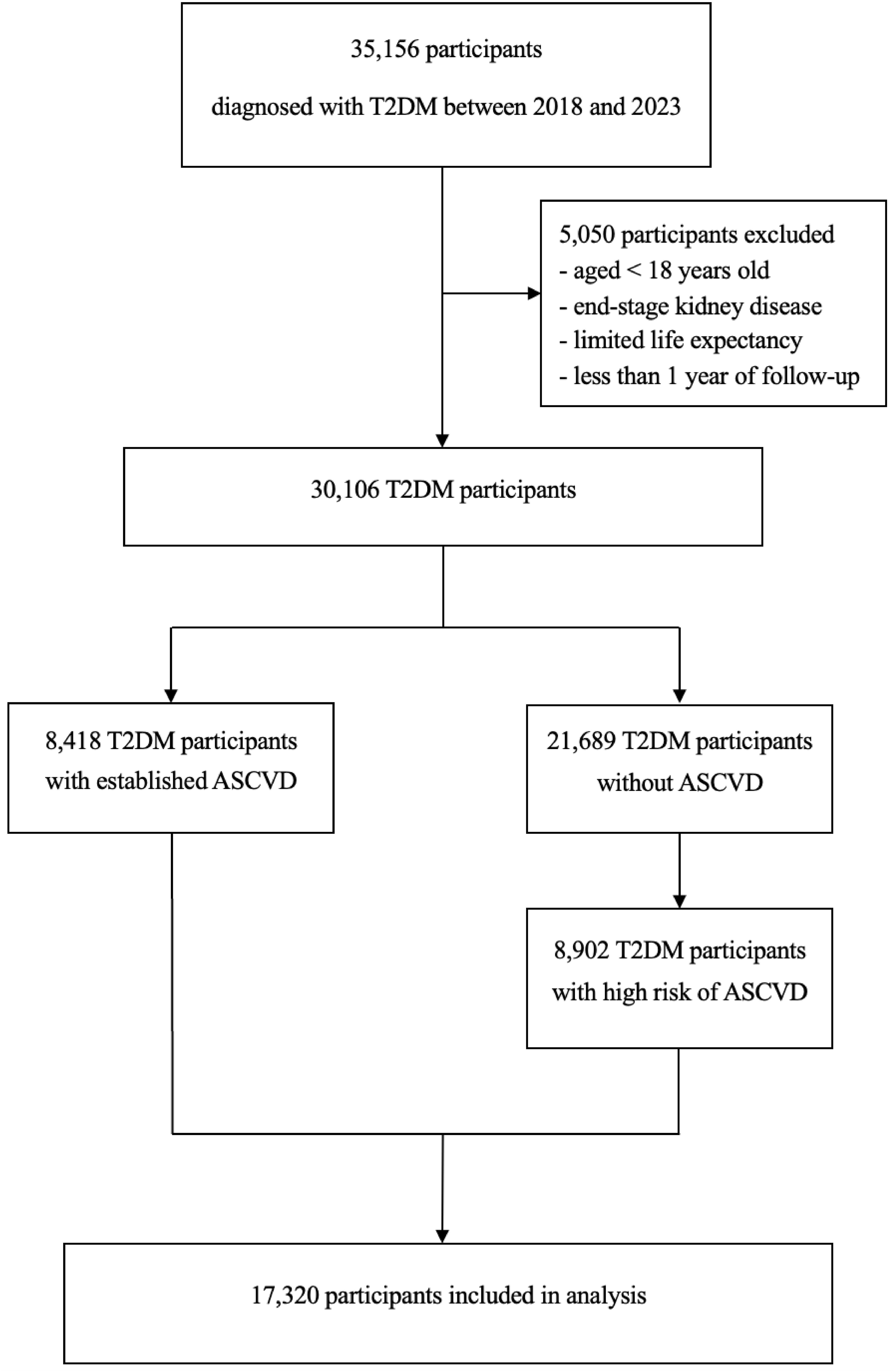
Trends in evidence-based therapy use among T2DM patients with established or high risk of ASCVD. Notes: ACEI, angiotensin-converting enzyme inhibitors; ARB, angiotensin-receptor blockers; GLP-1 RA, glucagon-like peptide-1 receptor agonists; SGLT-2i, sodium-glucose cotransporter-2 inhibitors.

### 3.4 Evidence-based Therapy

Compared with the overall cohort, patients prescribed GLP-1RA or SGLT2i were younger (mean age: GLP-1 RA: 61.1 years; SGLT2i: 63.0 years; overall: 66.3 years), had lower prevalence of heart failure (GLP-1 RA: 26.4%; SGLT2i: 21.4%; overall: 32.3%), had higher prevalence of dyslipidemia (GLP-1 RA: 87.2%; SGLT2i: 86.8%; overall: 81.5%), had fewer medical comorbidities (mean Charlson comorbidity index score: GLP-1 RA: 3.8; SGLT2i: 3.7; overall: 4.0) and had a lower percentage of patients with HbA1c < 7% (GLP-1 RA: 33.5%; SGLT2i: 36.6%; overall: 41.1%). Patients who prescribed a GLP-1 RA had higher prevalence of diabetes chronic complications (retinopathy: 16.1% vs 9.9%; neuropathy: 40.9% vs 27.6%) compared with the overall cohort (Table 3). The demographics and clinical characteristic of patients prescribed an ACEI or ARB, moderate-intensity statin and aspirin were displayed in Table 3.

**Table 3.**
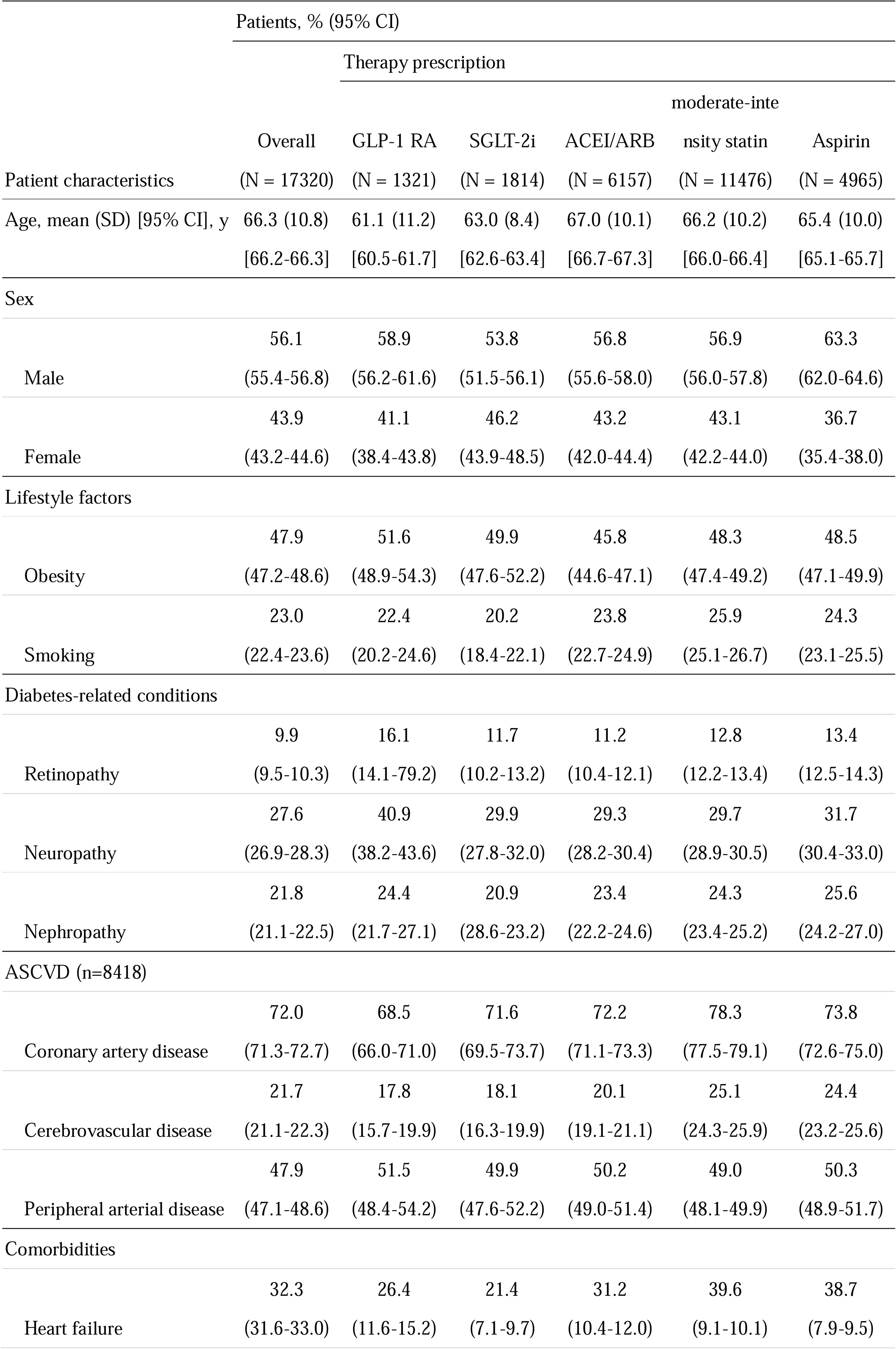
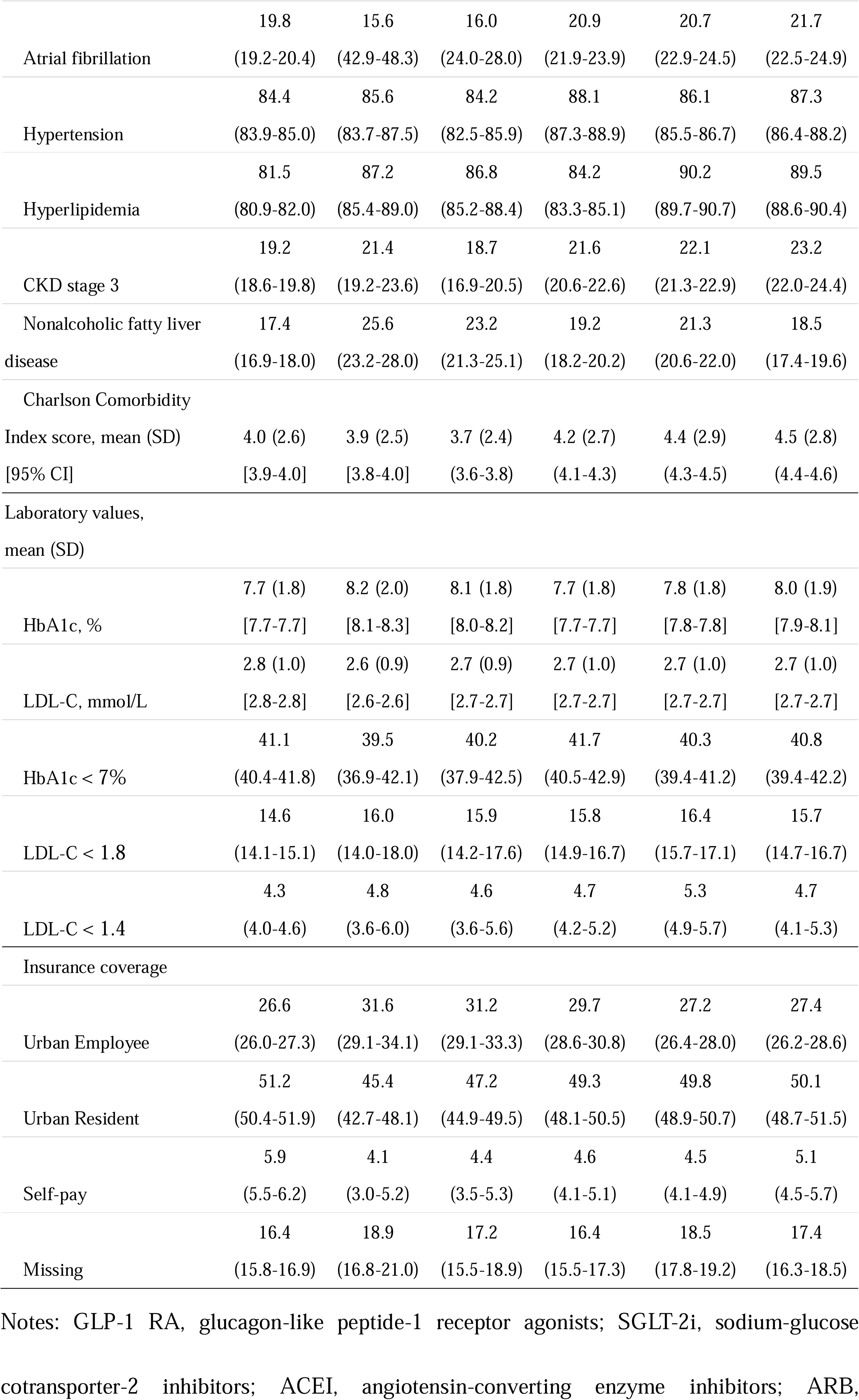
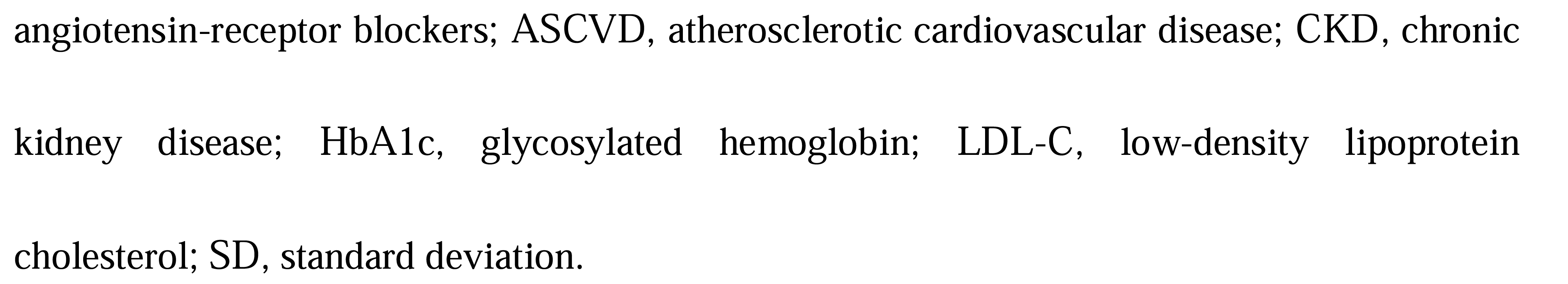
Patient characteristics at baseline by individual evident-based therapies.

## 4 Discussion

In our cohort of patients with T2DM and established or high risk of ASCVD, we demonstrated that the use of evidence-based cardiovascular preventive therapies was associated with a lower risk of 3-point MACE, HHF and end-stage kidney disease or doubling of serum creatinine level relative to non-use. And our study suggested that the overall usage of evidence-based cardiovascular preventive therapies between 2018 and 2023. The overall antihyperglycemic agents with proven cardiovascular benefits use was 7.6% for GLP-1 RA, and 10.5% for SGLT2i. The utilization rates were 35.5% for ACEI/ARB, 66.3% for moderate-intensity statins, and 28.7% for aspirin. Further, approximately half of patients received no evidence-based cardiovascular benefit medication.

To our knowledge, this study was one of the few to date to compare the cardiovascular and renal effectiveness of using or not-using evidence-based cardiovascular preventive therapies. Consistent with a meta-analysis of cardiovascular outcome trials, we found that benefits of GLP-1 RA, SGLT2i, ACEI/ARB, statins, and aspirin were observed for the outcomes of 3-point MACE, HHF and end-stage kidney disease or doubling of serum creatinine level [19–23].

The baseline of clinical characteristics was different between those using and not using evidence-based cardiovascular preventive therapies, and this might account for the failure to observe a difference in cardiovascular outcomes between the two groups. Consistent with a large cohort study, we observed that benefits for the outcomes of 3-point MACE, HHF, and end-stage kidney disease or doubling of serum creatinine level were significant difference between the two groups after adjusting for covariates[24].

Although both GLP-1 RA, SGLT-2i, ACEI/ARB, statins and aspirin have demonstrated cardiovascular benefits, the usage of these drugs is still low in the real-world [25,26]. Our finding suggested that the overall utilization of evidence-based cardiovascular preventive therapies was lower than expected, particularly for GLP-1 RA and SGLT2i. The findings that 14.0% of patients in our study prescribed either a GLP-1 RA or SGLT2i (GLP-1 RA: 7.6%, SGLT2i: 10.5%) are consistent with those of recent studies conducted in the US and Belgian [27–29]. Fortunately, our study found that the use of both GLP-1 RA and SGLT2i increased gradually from 2018 to 2023. However, as recently as 2023, two glucose-lowering therapies without proven cardiovascular benefit, namely alpha-glucosidase inhibitors and DPP-4i, continue to be used more frequently than either GLP-1 RA or SGLT2i.

Addressing these gaps in optimal pharmacotherapy is pivotal to improve patient outcomes. Reasons for continued underuse of these evidence-based therapies are likely multifactorial, and include the guidelines update, the relatively higher cost and out-of-pocket cost of new therapies, as well as clinical inertia. As we known, GLP-1 RA and SGLT2i both have demonstrated substantial benefits on major adverse cardiovascular events in those with T2DM and ASCVD [20,30]. Based on these data, the ADA has recommended GLP-1 RA and SGLT2i as first-line agents for patients with T2DM and ASCVD after metformin since 2019 [31]. In 2023, the ADA recommended that clinicians consider prescribing GLP-1 RA and SGLT2i prior to metformin for patients with T2DM and established or high risk of ASCVD [32]. In the wake of these updates, the use of GLP-1 RA and SGLT2i both slightly increased from 2018 through 2019, followed by a more than 3-fold from 2019 to 2023.

Despite the room for improvement, our findings support cautious optimism around the uptake of these agents in clinical practice. The use of GLP-1 RA rates doubled from 2020 through 2021, when the semaglutide was covered by health insurance in China. A similar trend was observed for SGLT2i, when the empagliflozin and dapagliflozin was covered by health insurance in 2020, the usage rates of these drugs doubled from 2019 to 2020. The rate of urban employee medical insurance was higher among those using evidence-based cardiovascular preventive therapies compared with those not using.

Therapeutic inertia within the realm of CVD prevention is a well-established barrier to widespread and consistent implementation of novel pharmacotherapeutic advances[33]. Herein, we observed slightly higher HbA1c levels and had a lower percentage of patients achieved HbA1c < 7% among those treated with GLP-1 RA and SGLT2i. These agents remained to be used less frequently than metformin or alpha-glucosidase inhibitors. This could imply that GLP-1 RA and SGLT2i were not considered as first-line therapeutic in T2DM individuals with high cardiovascular risk by clinicians. There are several strengths and limitations to this study. The primary strength of this study is that it was the first to compare the cardiovascular and renal effectiveness of using or not-using evidence-based cardiovascular preventive therapies. And we further confirmed the benefits of GLP-1 RA, SGLT-2i, ACEI/ARB, statins, and aspirin on the cardiovascular and renal outcomes for patients with T2DM and established or high risk of ASCVD. Second, we identified the rates of use of the evidence-based cardiovascular preventive therapies among T2DM patients with established or high risk of ASCVD. Additionally, we explored the possible reasons for the underuse of these evidence-based therapies. However, there are some limitations in this study. Firstly, given the nature of our study, it cannot prove causality or completely rule out residual confounding. We do our utmost to adjust for any possible confounding factors that may affect the primary outcomes. Secondly, our study sample consisting almost entirely of participants of Asian ethnicity, and whether the results can be extrapolated to non-Asian populations remains uncertain. Finally, our findings suggest that treatment discontinuation is common. However, the reason for patients switching from cardiovascular evidence-based therapies to other noncardiovascular agents was not illuminated in this study.

## 5 Conclusion

In conclusion, it is particularly concerning that relative to cardiovascular evidence-based therapies (GLP-1 RA, SGLT-2i, ACEI/ARB, statins, and aspirin), was associated with a lower risk of the 3-point MACE, HHF and end-stage kidney disease or doubling of serum creatinine level outcome in T2DM patients with ASCVD or high risk of ASCVD after adjusting for covariates. Despite estimates of evidence-based therapy prescription are considerably lower than expected, the rates of cardiovascular evidence-based therapies increased annually from 2018 to 2023. To continue this momentum, creative implementation science approaches will be necessary to further increase the use of these safe and effective medications in T2DM individuals with established or high risk of ASCVD.

## Data Availability

The administrative and clinical research databases used in this study are accessible to other researchers by contacting the corresponding author. The research data and data derivatives cannot be shared outside of the terms of these agreements.

## Acknowledgements

We would like to thank Natural Science Foundation of Xiamen Province, China for funding this work.

