## supplement for "Effectiveness and use of evidence-based cardiovascular preventive therapies in type 2 diabetes patients with established or high risk of atherosclerotic cardiovascular disease"

### eTable 1. The International Classification of Disease 10th Revision (ICD-10 code) used to identify diseases information.

| *Disease name* | *ICD-10 code* |
| --- | --- |
| Type 2 diabetes mellitus | E11, E11.0, E11.1, E11.2, E11.3, E11.4, E11.5, E11.6, E11.7, E11.8, E11.9 |
| Ischemic heart disease | I20.0, I20.1, I20.8, I20.9, I21.0, I21.1, I21.2, I21.3, I21.4, I21.9, I22.0, I22.1, I22.8, I22.9, I23.0, I23.1, I23.2, I23.3, I23.4, I23.5, I23.6, I23.8, I24.0, I24.1, I24.8, I24.9, I25.5, I25.1, I25.2, I25.3, I25.4, I25.5, I25.6, I25.8, I25.9 |
| Transient ischemic attacks | G45.0, G45.1, G45.2, G45.3, G45.4, G45.8, G45.9 |
| Stroke | I60.0, I60.1, I60.2, I60.3, I60.4, I60.5, I60.6, I60.7, I60.8, I60.9, I61.0, I61.1, I61.3, I61.4, I61.5, I61.6, I61.8, I61.9, I62.0, I62.1, I62.9, I63.0, I63.1, I63.2, I63.3, I63.4, I63.5, I63.6, I63.8, I63.9, I64.0, I69.0, I69.1, I69.2, I69.3,  I69.4, I69.8, |
| Peripheral arterial disease | I70.2, I73.9, I74.0, I74.2, I74.2, I74.3, I74.4, I74.5, I74.8, I74.9 |
| Obesity | E66.0, E66.1, E66.2, E66.8, E66.9 |
| Hypertension | I10, I11.0, I11.9, I12.0, I12.9, I13.9, I15.0, I15.1, I15.2, I15.8, I15.9 |
| Hyperlipidemia | E78.0, E78.1, E78.2, E78.3, E78.4, E78.5, E78.6, E78.8, E78.9 |

### eTable 2. Hazard Ratios of Use vs Non-use Evidence-based Cardiovascular Preventive Therapies Across Patient Subgroups.

|  | | Outcomes (subgroups) | | Group B Events (N) | | Group A  Events (N) | | Group B vs. Group A  HR (95% CI) | *p* |
| --- | --- | --- | --- | --- | --- | --- | --- | --- | --- |
|  | | **3-P MACE** | |  | |  | |  |  |
|  | | Male | | 134 (1501) | | 100 (1504) | | 0.95 (0.75-1.20) | 0.66 |
|  | | Female | | 94 (1109) | | 137 (1221) | | 0.99 (0.74-1.31) | 0.93 |
|  | | ASCVD | | 148 (1402) | | 153 (1391) | | 0.94 (0.75-1.18) | 0.59 |
|  | | High risk of ASCVD | | 80 (1208) | | 84 (1334) | | 0.99 (0.73-1.34) | 0.93 |
|  | | HF + | | 114 (935) | | 108 (835) | | 0.89 (0.69-1.16) | 0.39 |
|  | | HF - | | 114 (1675) | | 129 (1890) | | 0.95 (0.74-1.23) | 0.71 |
|  | | CKD + | | 100 (851) | | 96 (783) | | 0.90 (0.68-1.19) | 0.45 |
|  | | CKD - | | 128 (1759) | | 141 (1942) | | 0.98 (0.77-1.25) | 0.87 |
|  | **HHF** | | |  | |  | |  |  |
|  | Male | | | 42 (1501) | | 53 (1504) | | 0.80 (0.54-1.20) | 0.29 |
|  | Female | | | 37 (1109) | | 50 (1221) | | 0.91 (0.59-1.39) | 0.65 |
|  | ASCVD | | | 60 (1402) | | 76 (1391) | | 0.79 (0.56-1.11) | 0.17 |
|  | High risk of ASCVD | | | 19 (1208) | | 27 (1334) | | 0.85 (0.47-1.52) | 0.58 |
|  | HF + | | | 27 (935) | | 31 (835) | | 0.82 (0.49-1.34) | 0.46 |
|  | HF - | | | 52 (1675) | | 72 (1890) | | 0.85 (0.60-1.22) | 0.38 |
|  | CKD + | | | 49 (851) | | 58 (783) | | 0.81 (0.56-1.19) | 0.29 |
|  | CKD - | | | 30 (1759) | | 45 (1942) | | 0.79 (0.49-1.25) | 0.31 |
| **End-stage kidney disease, doubling of serum creatinine level** | | |  | |  | |  | |  |
| Male | | | 102 (1501) | | 125 (1504) | | 0.87 (0.67-1.14) | | 0.32 |
| Female | | | 96 (1109) | | 101 (1221) | | 0.96 (0.73-1.27) | | 0.78 |
| ASCVD | | | 93 (1402) | | 112 (1391) | | 0.87 (0.66-1.15) | | 0.32 |
| High risk of ASCVD | | | 105 (1208) | | 114 (1334) | | 0.94 (0.72-1.22) | | 0.64 |
| HF + | | | 122 (935) | | 133 (835) | | 0.89 (0.70-1.14) | | 0.36 |
| HF - | | | 76 (1675) | | 93 (1890) | | 0.87 (0.64-1.18) | | 0.38 |
| CKD + | | | 56 (851) | | 64 (783) | | 0.89 (0.62-1.28) | | 0.53 |
| CKD - | | | 142 (1759) | | 162 (1942) | | 0.92 (0.73-1.15) | | 0.46 |

Notes: group A (non-use of evidence-based cardiovascular preventive therapies); group B (use at least one evidence-based cardiovascular preventive therapy); 3-P MACE, 3-point major adverse cardiovascular event; HHF, hospitalization for heart failure; ASCVD, atherosclerotic cardiovascular disease; CKD, chronic kidney disease.

### eTable 3. Patient Characteristics by Overall Evidence-based Composite Score (Cohort 2).

| Patient characteristics | Evidence-based therapy score, % (95% CI) | |
| --- | --- | --- |
|  | 0 (n=7378) | 1-4 (n=9942) |
| Age, mean (SD) [95% CI], y | 67.1 (10.2) [67.0-67.2] | 65.7 (10.6) [65.6-65.8] |
| Sex |  |  |
| Male | 54.7 (53.5-55.9) | 57.1 (56.0-58.2) |
| Female | 45.3 (44.1-46.5) | 42.9 (41.8-44.0) |
| Lifestyle factors |  |  |
| Obesity | 46.8 (45.7-48.0) | 48.7 (47.6-49.7) |
| Smoking | 22.6 (21.5-23.7) | 23.3 (22.5-24.2) |
| Diabetes-related conditions |  |  |
| Retinopathy | 7.5 (6.8-8.2) | 11.7 (11.0-12.3) |
| Neuropathy | 23.2 (22.1-24.2) | 30.9 (30.0-31.8) |
| Nephropathy | 39.8(38.6-40.9) | 43.2 (42.2-44.2) |
| Diabetes treatment |  |  |
| Initiation of SGLT-2i or  GLP-1 RAs | 3.2 (2.7-3.6) | 6.7 (6.1-7.2) |
| Concurrent metformin | 62.3 (61.2-63.5) | 60.9 (59.9-61.9) |
| Concurrent second-generation  sulfonylurea | 26.3 (25.3-27.3) | 17.2 (16.4-17.9) |
| ASCVD (n=8418) |  |  |
| Coronary artery disease | 70.1 (69.0-71.1) | 73.4 (72.4-74.3) |
| Cerebrovascular disease | 19.2 (18.3-20.1) | 23.6 (22.7-24.4) |
| Peripheral arterial disease | 45.7 (44.6-46.9) | 49.5 (48.4-50.6) |
| Comorbidities |  |  |
| Heart failure | 29.3 (28.3-30.4) | 31.6 (30.6-32.5) |
| Atrial fibrillation | 20.1 (19.2-21.1) | 19.6 (18.7-20.4) |
| Hypertension | 80.6 (79.6-81.5) | 87.3 (86.5-88.2) |
| Hyperlipidemia | 75.2 (74.2-76.3) | 86.1 (85.3-86.8) |
| CKD stage 3 | 17.2 (16.3-18.1) | 20.7 (19.8-21.5) |
| Nonalcoholic fatty liver disease | 17.2 (16.2-18.1) | 17.6 (16.9-18.3) |
| Charlson Comorbidity Index score, mean (SD) [95% CI] | 3.8 (2.6) [3.8-3.8] | 4.1 (2.7) [4.1-4.1] |
| Laboratory values, mean (SD) |  |  |
| HbA1c, % | 7.7 (1.8) [7.7-7.7] | 7.8 (2.0) [7.8-7.8] |
| LDL-C, mmol/L | 2.8 (1.0) [2.8-2.8] | 2.7 (0.9) [2.7-2.7] |
| HbA1c < 7% | 41.3 (40.2-42.4) | 40.9 (39.9-41.9) |
| LDL-C < 1.8 | 14.9 (14.1-15.1) | 14.4 (15.9-20.1) |
| LDL-C < 1.4 | 4.2 (3.7-4.7) | 4.4 (4.0-4.9) |
| Insurance coverage |  |  |
| Urban Employee | 25.3 (24.3-26.4) | 27.6 (26.6-28.5) |
| Urban Resident | 54.3 (53.2-55.4) | 48.8 (47.8-49.8) |
| Self-pay | 5.2 (4.7-5.8) | 6.4 (5.9-6.9) |
| Missing | 15.3 (14.5-16.1) | 17.2 (16.5-17.9) |

Notes: SGLT-2i, sodium-glucose cotransporter-2 inhibitors; GLP-1 RA, glucagon-like peptide-1 receptor agonists; ASCVD, atherosclerotic cardiovascular disease; CKD, chronic kidney disease; HbA1c, glycosylated hemoglobin; LDL-C, low-density lipoprotein cholesterol; SD, standard deviation.

### eFigure 1. Trends in Oral Glucose-lowering Medication Use Among T2DM Patients with Established or High Risk of ASCVD, 2018-2023.

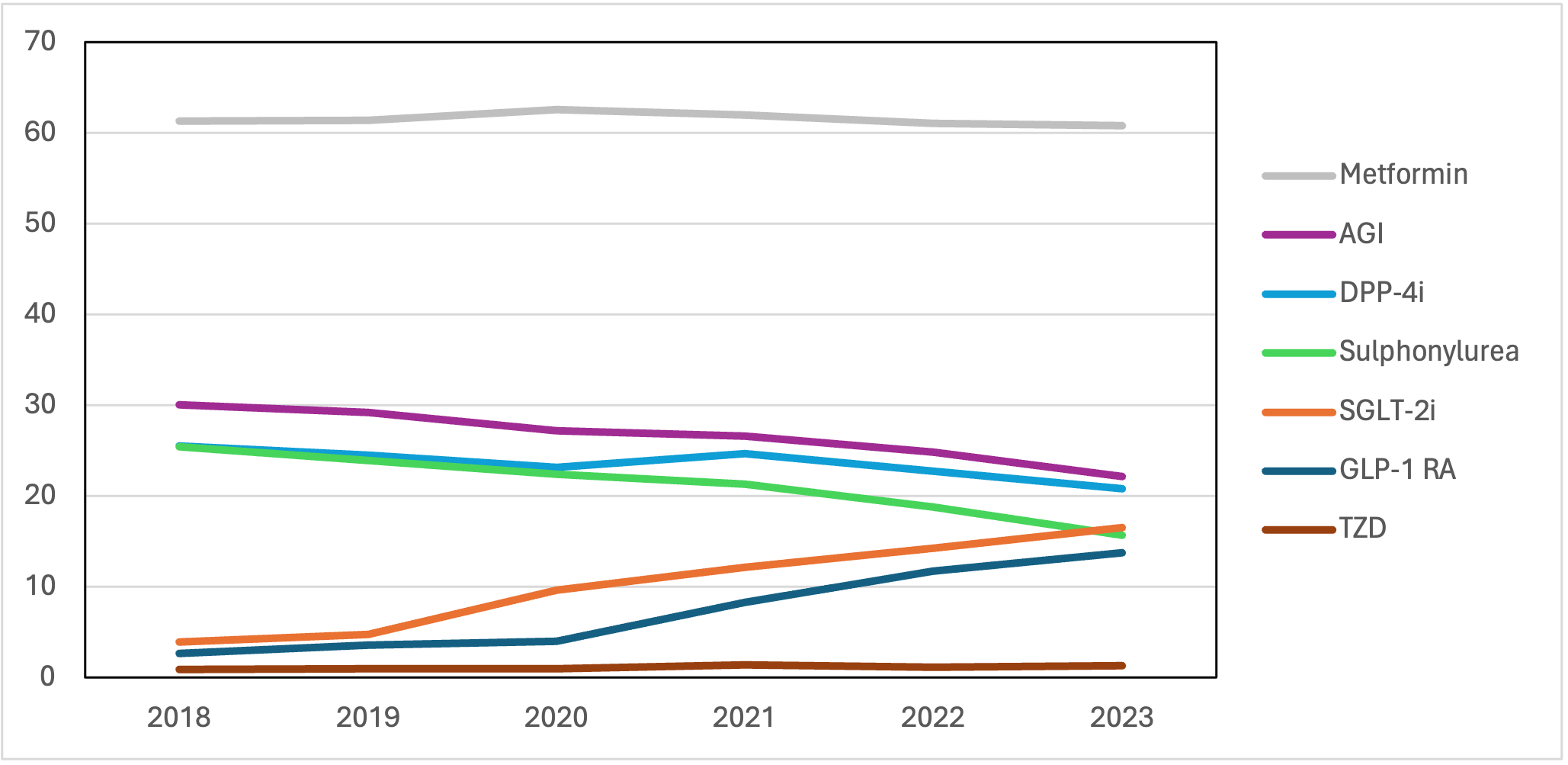

Notes: AGI, alpha-glucosidase inhibitors; DPP-4i, dipeptidyl peptidase-4 inhibitors; SGLT-2i, sodium-glucose cotransporter-2 inhibitors; GLP-1 RA, glucagon-like peptide-1 receptor agonists; TZD, thiazolidinediones.

### eFigure 2. Trends in Blood Pressure-lowering Medication Use Among T2DM Patients with Established or High Risk of ASCVD, 2018-2023.

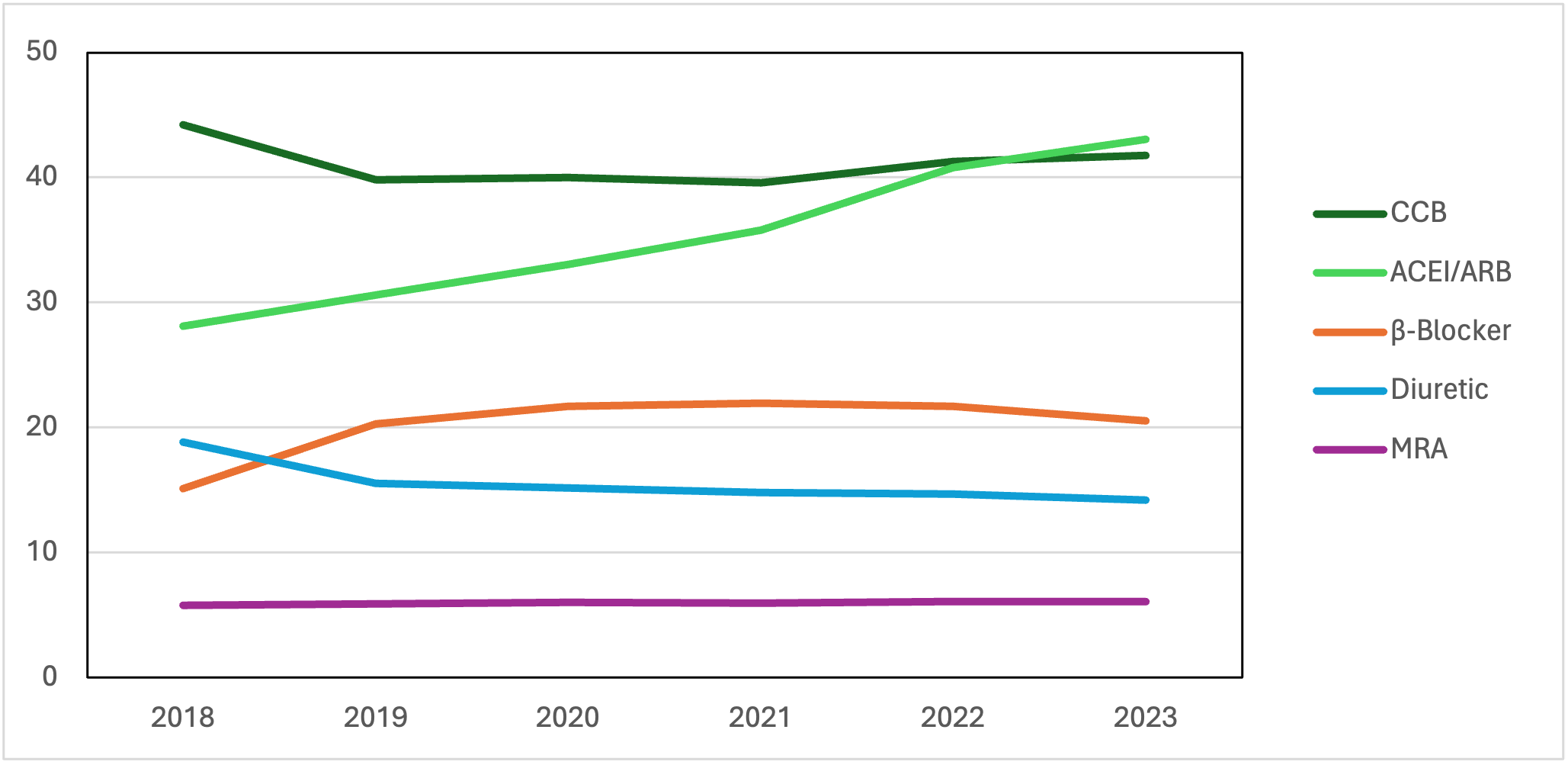

Notes: CCB, calcium channel blockers; ACEI, angiotensin-converting enzyme inhibitors; ARB, angiotensin-receptor blockers; MRA, mineralocorticoid receptor antagonists.

### eFigure 3. Trends in Lipid-lowering Medication Use Among T2DM Patients with Established or High Risk of ASCVD, 2018-2023.

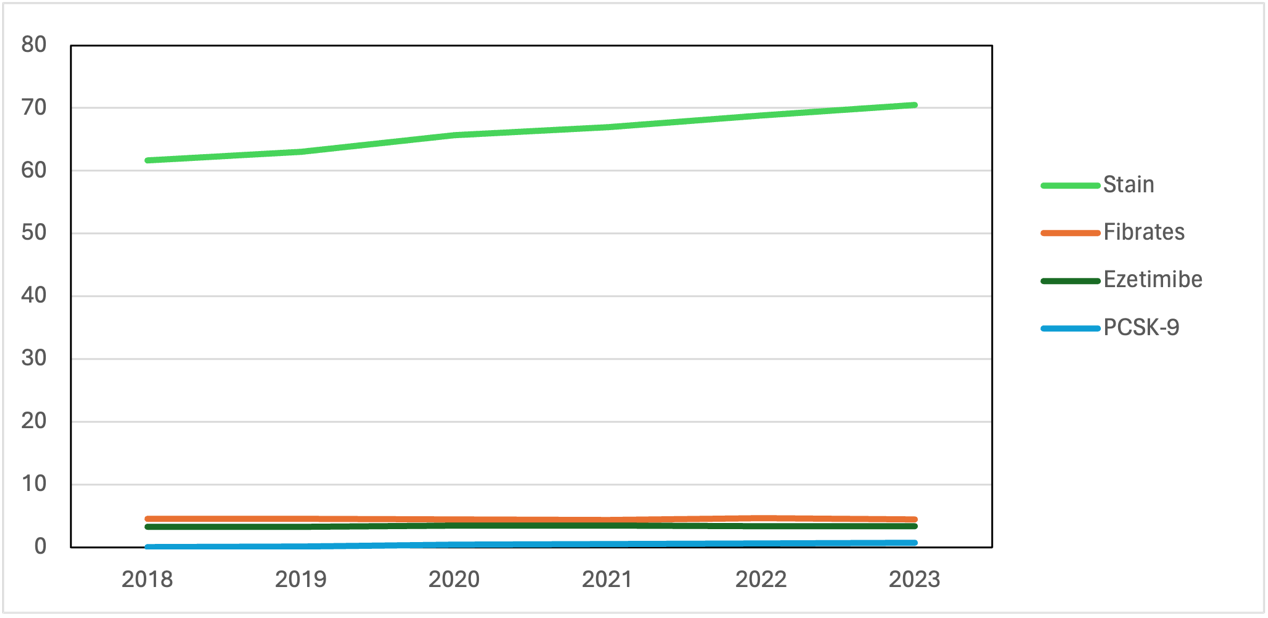

Notes: PCSK-9 proprotein convertase subtilisin/kexin type-9.

### eFigure 4. Use of Evidence-based Cardiovascular Preventive Therapies in T2DM Patients with Established or High Risk of ASCVD, 2018-2023.

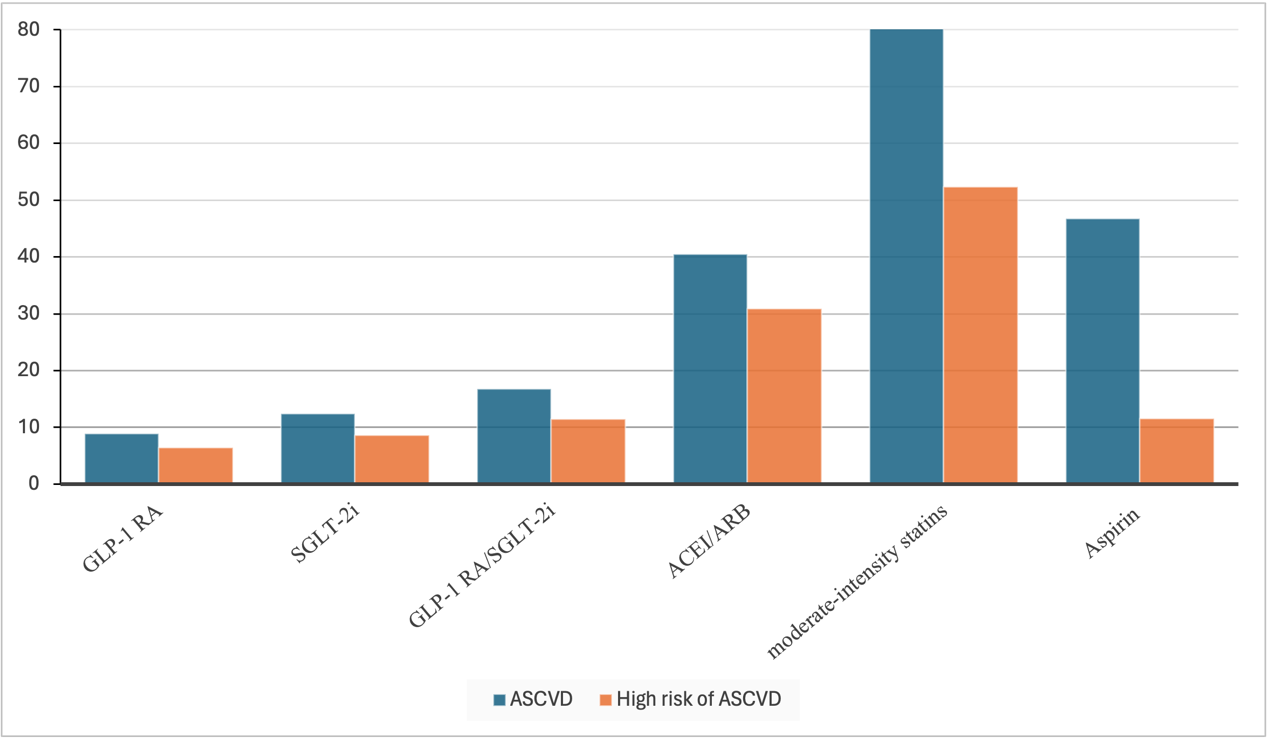

Notes: GLP-1 RA, glucagon-like peptide-1 receptor agonists; SGLT-2i, sodium-glucose cotransporter-2 inhibitors; ACEI, angiotensin-converting enzyme inhibitors; ARB, angiotensin-receptor blockers; ASCVD, atherosclerotic cardiovascular disease.
